# Genome-wide meta-analysis of iron status biomarkers and the effect of iron on all-cause mortality in HUNT

**DOI:** 10.1101/2021.09.20.21262960

**Authors:** Marta R Moksnes, Ailin Falkmo Hansen, Sarah E Graham, Sarah A Gagliano Taliun, Kuan-Han Wu, Wei Zhou, Ketil Thorstensen, Lars G Fritsche, Dipender Gill, Amy Mason, Francesco Cucca, David Schlessinger, Gonçalo R. Abecasis, Stephen Burgess, Bjørn Olav Åsvold, Jonas B Nielsen, Kristian Hveem, Cristen J Willer, Ben M Brumpton

**Affiliations:** K.G. Jebsen Center for Genetic Epidemiology, Department of Public Health and Nursing, Faculty of Medicine and Health Sciences, NTNU – Norwegian University of Science and Technology, Trondheim, Norway; Division of Cardiovascular Medicine, Department of Internal Medicine, University of Michigan, Ann Arbor, MI, USA; Faculty of Medicine, University of Montréal, Montréal, QC H3T 1J4, Canada; Montréal Heart Institute, Montréal, QC H1T 1C8, Canada; Department of Computational Medicine and Bioinformatics, University of Michigan, Ann Arbor, MI, USA; Analytic and Translational Genetics Unit, Department of Medicine, Massachusetts General Hospital, Boston, MA, USA; Stanley Center for Psychiatric Research, Broad Institute of MIT and Harvard, Cambridge, MA, USA; Department of Clinical Chemistry, St. Olavs hospital Trondheim University Hospital, Trondheim, Norway; Department of Biostatistics, University of Michigan School of Public Health, Ann Arbor, MI, USA; Center for Statistical Genetics, University of Michigan School of Public Health, Ann Arbor, MI, USA; Department of Epidemiology and Biostatistics, School of Public Health, Imperial College London, London, UK; Clinical Pharmacology and Therapeutics Section, Institute for Infection and Immunity, St George’s, University of London, London, UK; Clinical Pharmacology Group, Pharmacy and Medicines Directorate, St George’s University Hospitals NHS Foundation Trust, London, UK; Novo Nordisk Research Centre Oxford, Old Road Campus, Oxford, UK; Department of Public Health and Primary Care, University of Cambridge, Cambridge, UK; National Institute for Health Research Cambridge Biomedical Research Centre, University of Cambridge and Cambridge University Hospitals, Cambridge, UK; Istituto di Ricerca Genetica e Biomedica, Consiglio Nazionale delle Ricerche (CNR), Cagliari, Italy; Dipartimento di Scienze Biomediche, Università degli Studi di Sassari, Sassari, Italy; Laboratory of Genetics, National Institute on Aging, US National Institutes of Health, Baltimore, Maryland; Medical Research Council Biostatistics Unit, University of Cambridge, Cambridge, UK; Department of Endocrinology, Clinic of Medicine, St. Olavs hospital Trondheim University Hospital, Trondheim, Norway; HUNT Research Centre, Department of Public Health and Nursing, NTNU – Norwegian University of Science and Technology, Levanger, Norway; Department of Epidemiology Research, Statens Serum Institute, Copenhagen, Denmark; Department of Cardiology, Copenhagen University Hospital, Copenhagen, Denmark; Department of Human Genetics, University of Michigan, Ann Arbor, MI, USA; Clinic of Medicine, St. Olavs hospital Trondheim University Hospital, Trondheim, Norway

## Abstract

Iron is essential for many biological processes, but iron levels must be tightly regulated to avoid harmful effects of both iron deficiency and overload. Here, we perform genome-wide association studies on four iron related biomarkers (serum iron, serum ferritin, transferrin saturation, total iron binding capacity) in the Trøndelag Health Study (HUNT), the Michigan Genomics Initiative (MGI) and the SardiNIA study, followed by their meta-analysis with publicly available summary statistics, analyzing up to 257 953 individuals. We identify 127 genetic loci associated with iron traits. Among 19 novel protein-altering variants, we observe a rare missense variant (rs367731784) in HUNT, which suggests a role for *DNAJC13* in transferrin recycling. We further validate the latest genetic risk scores for each biomarker in HUNT (6% variance in serum iron explained) and present linear and non-linear Mendelian randomization analyses of the traits on all-cause mortality. We find evidence of a harmful effect of increased serum iron and transferrin saturation in linear analyses that estimate population-averaged effects. However, there was weak evidence of a protective effect of increasing serum iron at the very low end of its distribution. Our findings contribute to our understanding of the genes affecting iron status and its consequences on human health.

## Introduction

Iron is essential for a variety of physiological processes in the human body, but excess iron is toxic. Iron overload is associated with a wide range of health problems, including liver damage, type 2 diabetes, cardiovascular disease and neurodegenerative diseases such as Alzheimer’s disease^1–3^, while long-term iron deficiency causes anemia, which can disrupt cognitive function and the immune system^4–6^. Because of the damaging effects of both deficiency and overload, iron metabolism is tightly regulated^7^.

Iron is bound, transported and delivered around the body by the transferrin glycoprotein^8^, while the main intracellular iron storage, ferritin, provides a long-term reserve of iron for formation of hemoglobin and other heme proteins^9–11^. Serum iron, serum ferritin, transferrin saturation percentage (TSP) and the total iron binding capacity (TIBC) of transferrin are biochemical measurements that are commonly used together to assess an individual’s iron status^12^.

Mutations in various iron metabolism genes can cause both iron deficiency and overload^13–15^. Genetic variants in the transferrin gene, *TF*, and in the homeostatic iron regulator gene, *HFE*, have been estimated to account for about 40% of genetic variation in transferrin levels^16^. A recent genome-wide association study (GWAS) meta-analysis^17^ of serum iron, ferritin, TSP and TIBC from Iceland, UK and Denmark reported 46 novel loci associated with at least one of these biomarkers, implicating proteins involved in iron homeostasis. Identifying additional genetic loci associated with iron status could further increase our understanding of pathomechanisms underlying dysregulated iron levels. Furthermore, genetic variants from the most recent study^17^ could improve existing genetic risk scores (GRS) that have been widely used to assess the causal associations of iron status on a range of outcomes using Mendelian Randomization^18–23^ (MR). However, the new GRSs have not yet been validated in an independent study. Further, despite the observed damaging effects of both very high and very low iron stores, no previous MR studies have investigated the shape of the associations between genetically proxied iron status biomarkers and mortality. By validating the most recent genetic risk scores and using MR in an independent study (HUNT), we provide robust and novel insights into the causal associations between iron status biomarkers and all-cause mortality, particularly regarding non-linear relationships.

To discover novel genetic variants associated with iron status, we combine three approaches: (i) genome-wide association studies of variants deeply imputed from the TOPMed reference panel^24^ in the Trøndelag Health Study (HUNT)^25^ and the Michigan Genomics Initiative (MGI), as well as variants imputed from a cohort specific reference panel in SardiNIA^26^ (ii) association tests with genotyped coding variants selected from low-coverage (5x) whole-genome sequencing, (iii) genome-wide meta-analyses of HUNT, MGI, SardiNIA and summary statistics from deCODE, Interval and the Danish Blood Donor Study (DBDS)^17^. The analyses included up to 257 953 individuals (57% females, 43% males) with measured iron status biomarkers. We evaluate the variance explained by previously published variants for serum iron, serum ferritin, TSP and TIBC in HUNT. Furthermore, we use the genetic variants for the iron status biomarkers to estimate the average causal effect of a population shift in the biomarker distributions on all-cause mortality (the population-averaged effect), and for the first time, investigate the shape of the causal relationships using non-linear Mendelian Randomization.

## Results

### Discovery of genetic loci associated with iron status

We identified 127 genetic loci (82 novel for at least one trait) associated (p-value < 5×10^−8^) with the four iron traits, serum iron, serum ferritin, TIBC and TSP, (Supplemental Table 1, Supplemental Figures 1-4) in genome-wide association meta-analyses of the iron status biomarkers in 6 cohorts: HUNT, MGI, SardiNIA, deCODE, Interval and DBDS (Supplemental Table 2). Among the 77 unique index variants (the variants with the lowest p-value) in novel loci that had been imputed in more than one study, 60 had consistent directions of effects across all the analyzed studies. We also identified three novel missense variants associated with at least one iron trait among the variants selected for targeted genotyping in HUNT (Supplemental Table 3).

Genes in several associated loci coded for proteins with established functions in iron homeostasis (*TF* [transferrin]) *SLC25A37* [mitoferrin-1], *SLC25A28* [mitoferrin-2], *SLC11A2* [divalent metal-transporter 1] and *SLC40A1* [ferroportin-1], *HFE* [homeostatic iron regulator], *TFRC* [transferrin receptor], *TFR2* [transferrin receptor 2], *HAMP* [hepcidin], *ERFE* [erythroferrone], *HMOX1* [heme oxygenase], *IREB2* [iron responsive element binding protein 2], *EPAS1* [endothelial PAS Domain Protein 1] and *TMPRSS6* [transmembrane serine protease 6])^7,27–29^ Four of these loci (*SLC25A28, HMOX1, IREB2, EPAS1*) had not been reported for iron status biomarkers in GWAS studies before, providing additional confidence in the GWAS we report. With two exceptions (*HAMP, TFR2*), these genes were the nearest gene to the index variant in the locus.

### Protein-altering variants in meta-analysis loci

We identified 32 protein-altering single nucleotide polymorphisms (SNPs) in the meta-analysis, which were either index variants or variants in strong linkage disequilibrium (LD) (R^2^>0.8 or D’=1.0) with an index variant (Supplemental Table 1). In addition to SNPs known to be related to diseases such as hemochromatosis, atransferrinemia and iron deficiency anemia^15,30–34^, and variants that had previously been reported for at least one of the iron traits^17,35^, we identified 11 protein-altering variants in novel iron status loci (Supplemental Table 4): rs9427398 (*FCGR2A*), rs2437150 (*SPRTN*), rs1047891 (*CPS1*), rs41274050 (*A1CF*), rs1935 (*JMJD1C*), rs3742049 (*COQ5*), rs4149056 (*SLCO1B1*), rs2070863 (*SERPINF2*), rs883541 (*WIPI1*), rs1800961 (*HNF4A*) and rs738409 (*PNPLA3*).). In known iron status loci, we further identified eight protein-altering variants not previously reported for any iron traits: rs367731784 (*DNAJC13*), rs3812594 (*SEC16A*), rs34376913 (C9orf163), rs445520 (*SLC11A2*), rs28929474 (*SERPINA1*), rs737700 (*C16orf71*), rs77542162 (*ABCA6*) and rs34654230 (*RCN3*).

### Custom genotyped variants in HUNT: Protein-altering variants in iron status loci

Among the targeted candidate variants in HUNT identified by sequencing and clinical studies, we identified three additional, novel protein-altering variants (Supplemental Table 3) that were not included in the meta-analyses, and which were associated with iron status biomarkers. These were located in *NRM* (rs374815811), *HLA*-*DRB5* (rs701884) and *TFR2* (chr7:100629337:A:T, GRCh38).

### Heritability and genetic correlation of iron status markers

We estimated the respective narrow-sense SNP heritability (variance explained, V_g_/V_p_ ± 1SE) of serum iron (0.15± 0.01), TIBC (0.43 ± 0.01) and TSP (0.21±0.01) in HUNT using genome-wide complex trait analysis (GCTA)^36^. We found pair-wise genetic correlations between 11% and 75% (Supplemental Table 5) for the four iron status biomarkers using LD Score Regression (LDSC)^37^ with the meta-analysis summary statistics. The TSP phenotype was derived from the serum iron and TIBC measurements, giving rise to the two strongest genetic correlations. The weakest correlation (iron vs TIBC) did not reach nominal significance (p-value=0.35).

### Functional mapping

We used Bayesian colocalization analysis to identify 94 unique pairs of GWAS loci and cis-expression quantitative trait locus (cis-eQTL) signals that showed sufficient overlap in at least one tissue to be consistent with a shared causal variant for the gene expression and the iron status biomarker (Supplemental Table 6). We found associations in a range of tissues which highlighted genes with established roles in iron metabolism (*TF* [posterior probability of a common causal variant, PP4=0.96], *TMPRSS6* [PP4=0.82], *ERFE* [PP4=0.97-0.98], *IREB2* [PP4=0.80], *SLC40A1* [PP4=0.79-0.96])^13,27^. Additionally, our results confirmed previously reported genes (*DUOX2* [PP4=0.76], *HBS1L* [PP4=0.98], *IL6R* [PP4=0.81-0.82], *SLC25A37* [PP4=0.85], *ABO* [PP4=0.97], *RNF43* [PP4=0.99])^17,35^, and identified novel genes interacting with previously reported genes, for example *DUOXA2*^38^. Several iron status loci were also colocalized with cis-eQTL signals for genes in the major histocompatibility complex (MHC) other than *HFE*^39^, as well as with transcription regulators^40–42^, additional transporter proteins^43,44^ and transferases^45,46^.

Using Data-driven Expression Prioritized Integration for Complex Traits (DEPICT)^47^ we found an enriched (false discovery rate [FDR] <0.05) expression of ferritin associated genes in the urogenital system, digestive system, and the hemic and immune system (Supplemental Table 7). Serum iron, TSP and TIBC associated genes were not enriched in any tissue types at FDR<0.05, however the strongest enrichment for genes in all three traits were found in liver tissue, and particularly in hepatocytes (TSP, TIBC). The top ten genes per trait when prioritized based on similarity between the associated (p-value < 5×10^−8^) loci, included known iron regulatory genes (*TFR2, HAMP, TFRC, SLC40A1*), genes in which we had identified protein-altering variants (*IL6R, F5, GCKR, DUOX2, SERPINA1, ABCA6, SLCO1B1*), genes we found in the colocalization analysis (*DUOXA2, IL1RN, SLC25A37*), as well as genes predicted to have iron ion binding and heme binding properties in gene ontology analyses (*CYP3A43, CYP3A5*)^48^ (Supplemental Table 8). Finally, we used DEPICT and found gene sets enriched with iron status associated genes (Supplemental Table 9). Most of the top ten gene sets at FDR<0.05 were from iron and TSP associated loci: Several were related to the liver (including abnormal liver physiology and gene sets related to metabolic processes), but also to inflammation (acute-phase response, decreased leukocyte cell number), coagulation (coagulation factor protein-protein interaction networks) and neurodevelopment (abnormal myelination). Most of the top ten gene sets enriched with ferritin and TIBC associated loci did not reach FDR<0.05, but included decreased circulating iron levels, decreased spleen iron level, gene sets related to red blood cells (decreased hemoglobin, decreased hematocrit, erythrocyte homeostasis and differentiation), as well as liver fibrosis and liver inflammation.

We identified the 1% top ranked genes per trait based on both physical distance to the associated genetic variants and functional similarity to other associated genes (Supplemental Tables 10-13) using Polygenic Priority Scores (PoPS)^49^. The prioritized genes included the main known iron regulatory genes, several genes that were nearest to the meta-analysis index variants and novel genes in which we identified protein-altering variants (*WIPI1, SERPINF2, HNF4A*), further supporting a role for these genes in iron biology.

### Phenome-wide association study (PheWAS) of biomarker loci

In total, 69 of the meta-analysis index variants were significantly associated (p-value < 2.4×10^−7^) with at least one additional phenotype (‘phecode’^50^), blood biomarker or continuous trait in UK Biobank, and 97 phenotypes were significantly associated with at least one variant (Supplemental Table 14). The associations spanned numerous biological domains, but most associations were within the endocrine/metabolic (288 variant-trait associations), digestive (139 variant-trait associations) and genitourinary (46 variant-trait associations) domains. The strongest associations (p-values < 1×10^−300^) were within the hematopoietic, digestive and endocrine/metabolic domains: The *HFE* variants rs1800562, rs144861591 and rs79220007 were associated with disorders of mineral metabolism, and in particular with disorders of iron metabolism, the *SLCO1B1* variant rs2900478 was associated with bilirubin, the *ASGR2*;*ASGR1* variant rs186021206 was associated with alkaline phosphatase, and the *GCKR* variants rs1260326 and rs11336847 were associated with triglycerides. Overall, all the GRSs for the four iron status biomarkers were associated with disorders of mineral metabolism, in particular iron metabolism (Figure 1, Supplemental Table 15). Several GRSs were associated with anemias (iron, ferritin, TSP) and with coagulation defects (ferritin, TSP). Finally, the GRS for TIBC was also associated with liver cirrhosis without mention of alcohol, and the GRS for ferritin was associated with chronic non-alcoholic liver disease, and with phlebitis and thrombophlebitis.

**Figure 1:**
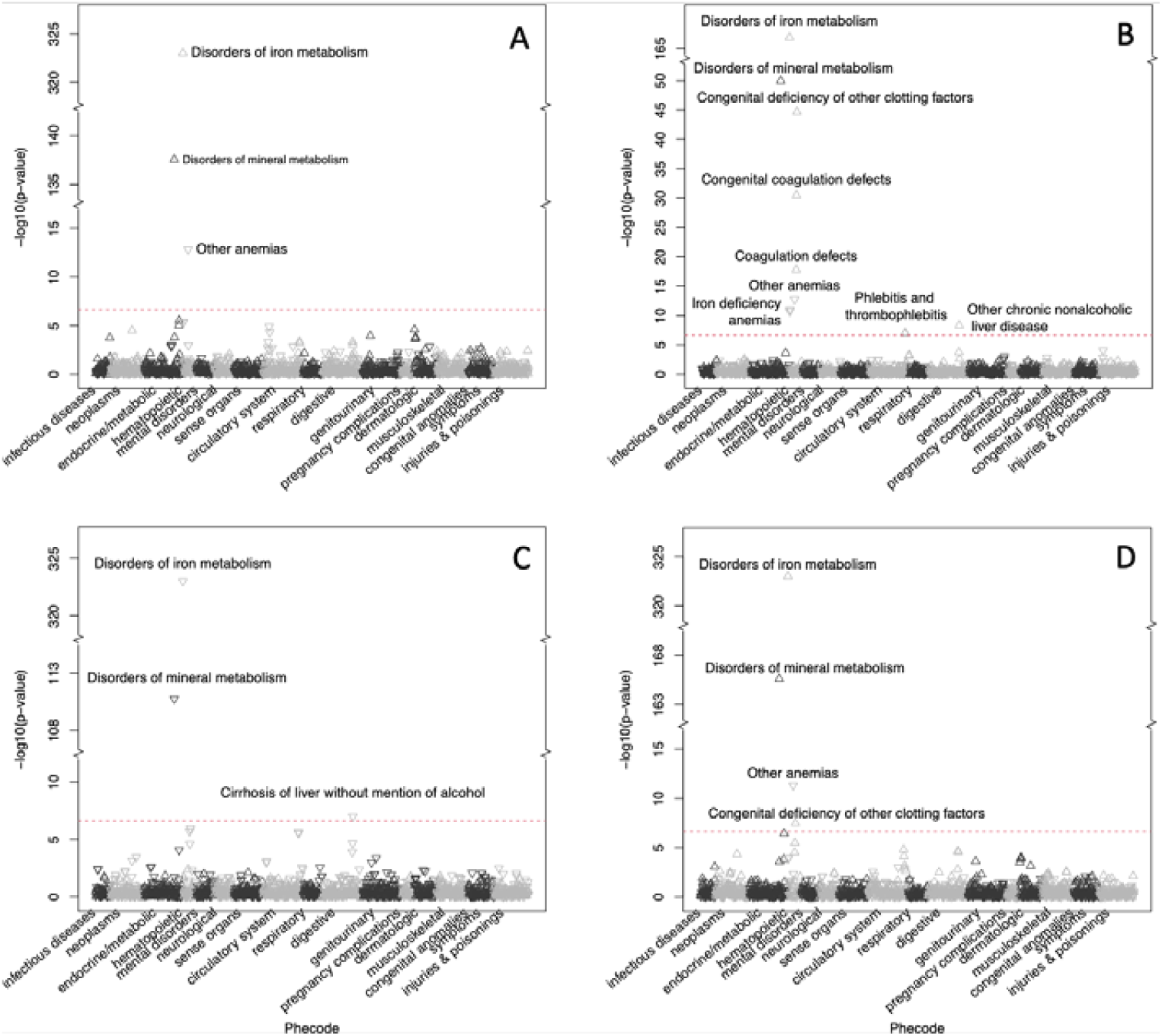
GRS-PheWAS: Phenome-wide associations between the GRS for each biomarker (serum iron [A], serum ferritin [B], total iron binding capacity [C] and transferrin saturation percentage [D]) and 1 688 phecodes, blood biomarkers and continuous traits in the UK Biobank. Triangles pointing upwards indicate a positive association between the phenotype and the GRS (where a higher GRS score represents higher level of the iron status biomarker) and vice versa. Associations with p-values < 10^−322^ are plotted at 10^−322^. The Bonferroni corrected p-value cut-off (2.4×10^−7^) is given as a red dotted line.

### Linear Mendelian Randomization

The linear MR (ratio of coefficients method) indicated an increased mortality risk with increased serum iron and TSP, with the point estimates suggesting that an increase of 1 standard deviation (SD) in both serum iron (1 SD = 6.3µmol/L) and TSP (1 SD = 11.3 percentage points) would lead to an increased risk of mortality of 7% (Table 1). The estimates for ferritin and TIBC were not statistically significant, however the point estimates for a 1 standard deviation increase in serum ferritin (1 SD = 46µg/L) and TIBC (1 SD = 9.2 µmol/L) were a 4% increase and 3% decrease in mortality, respectively (Table 1). The estimate for ferritin was also very imprecise.

**Table 1:** Linear Mendelian randomization ratio of coefficient estimates: Hazard ratios with 95% confidence intervals for all-cause mortality are given per 1 standard deviation increase in the biomarker. The sample size is given as N.

| Biomarker | N | Hazard ratio (95% CI) | P-value |
| --- | --- | --- | --- |
| Serum iron | 56 654 | 1.07 (1.01 to 1.14) | 0.03 |
| Serum ferritin* | 2 335 | 1.04 (0.26 to 4.22) | 0.95 |
| TSP | 56 651 | 1.07 (1.02 to 1.12) | 0.01 |
| TIBC | 56 654 | 0.97 (0.93 to 1.01) | 0.13 |

Confidence interval (CI), Transferrin saturation percentage (TSP), Total iron binding capacity (TIBC)
\*Non-pregnant women, 20-55 years old

### Non-linear Mendelian Randomization

To investigate a potential non-linear causal association between iron status and all-cause mortality, we used the GRSs as instruments for serum iron (F-statistic=3618, R^2^=0.06), TIBC (F-statistic=8373, R^2^=0.129), TSP (F-statistic=6811, R^2^=0.107) and ferritin (F-statistic=37.81, R^2^=0.015) in a non-linear Mendelian randomization analysis and estimated the shape of the associations between the genetically predicted traits and all-cause mortality (Figure 2). The median follow-up time was 23.6 years. After performing a statistical test for whether the best-fitting non-linear model of degree 1 fitted the data better than a linear model (p-values: 0.60, 0.05, 0.23 and 1 for iron, ferritin, TIBC and TSP respectively), generally, we did not find strong statistical evidence supporting a non-linear relationship over a linear one for the associations between any of the genetically proxied iron traits and all-cause mortality. However, the point estimates for serum iron do follow a J-shape, with a negative slope at very low levels of serum iron and a constant positive slope above 10 µmol/L. The point estimates were however imprecise at the tails of the distribution. The other analyses indicate a lower risk at higher TIBC and lower TSP and ferritin levels, with a weak indication (p-value=0.05) of a non-linear effect for ferritin. Post-hoc sensitivity analyses using genetic instruments that were consistent with systemic iron status (increased iron, ferritin, and TSP, and decreased TIBC) rather than just representing a single biomarker, gave similar results (Supplemental Figure 5).

**Figure 2:**
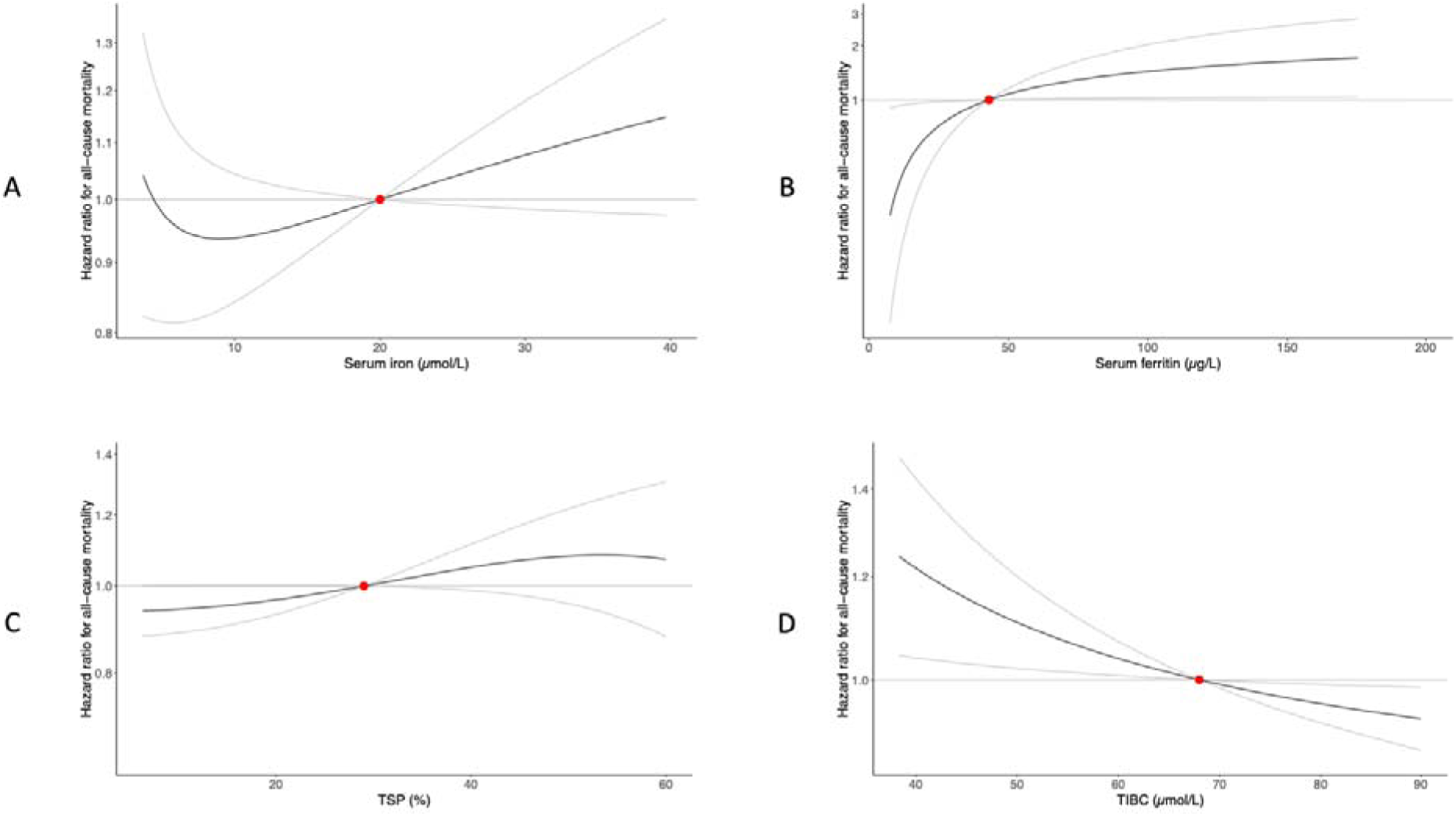
Non-linear Mendelian randomization: Dose-response curves (black) between iron traits and all-cause mortality in HUNT (gray lines give 95% confidence interval). The x-axis gives A: serum iron levels (µmol/L), B: serum ferritin (µg/L), C: transferrin saturation (%) and D: total iron binding capacity (TIBC) (µmol/L). The y-axis gives the hazard ratios for all-cause mortality with respect to the reference values (red dot), which represent the established target values (iron, TIBC, TSP)^51^ or median value (ferritin) for the traits. The curve gradients represent the localized average causal effect at each point.

## Discussion

We performed the largest GWAS meta-analysis to-date of iron status biomarkers and identified 127 genetic loci (82 novel) associated with iron status, including 19 novel protein-altering variants. Although 82 loci were classified as novel for at least one of the tested iron traits, some had known associations with other tested traits or with related traits such as hemoglobin levels and red blood cell count and volume^52^. Because iron traits are biologically linked, we might expect to find the same loci associated with several of the tested traits, as seen for loci with established roles in iron metabolism (e.g. *HFE, TF* and *TMPRSS6*) and others. We confirmed the genetic similarity between iron status biomarkers by observing a genetic correlation of 75% between serum iron and TSP. However, most of the loci were specifically associated with a single trait, which was in line with the low genetic correlation between other pairs of biomarkers.

Overall, our findings are consistent with established knowledge about iron homeostasis and the role of iron in various biological processes: The PheWAS analyses linked the meta-analysis loci to many different traits and phenotypes, particularly within the hematopoietic, digestive, endocrine/metabolic and genitourinary domains. Further, in four novel loci, the genes nearest to the index variant encoded proteins with established roles in iron regulation: 1) the mitochondrial iron transporter mitoferrin-2 (*SLC25A28*), 2) heme oxygenase 1 (*HMOX1*), which catalyzes heme degradation, 3) the iron-responsive element binding 2 (*IREB2*), and 4) *PAS1*, which regulates erythropoiesis according to cellular iron availability^28^. Further, the novel protein-altering variants were found in genes associated with various biological traits and functions, which potentially highlight the many biological processes both involved in and dependent on iron and iron regulation. These included, but were not limited to, genes involved in or associated with: 1) Iron gut absorption, regulation and transport (*TFR2, SLC11A2, DNAJC13*)^8,54,53^, where we found a rare (minor allele frequency (MAF)=0.0009) protein-altering variant with moderate effect size in *DNAJC13*, a gene suggested to be involved in transferrin recycling^53^. This variant was only imputed in HUNT, where it was more than 100 times more common than in other non-Finnish Europeans (https://gnomad.broadinstitute.org/variant/rs367731784)^55^ ; 2) Concentrations of hemoglobin (*SPRTN, FCGR2A, CPS1, PNPLA3, ABCA6*)^49,56–58^, which holds more than two thirds of the body’s iron^1^, and bilirubin (*SLCO1B1*)^59^, which together with iron are products of heme degradation; 3) Liver-related traits (*ABCA6, HNF4A, PNPLA3*)^60,61^; 4) Iron-dependent (putative) tumor suppression (*JMJD1C*)^62^; 5) Iron accumulation in the brain (*WIP1*, associated via its homolog *WIPI4*^63^) and 6) Coagulation and immune response (*SERPINA1, SERPINF2*)^64^. A previous study also identified a different protein-altering SNP in the serine protease inhibitor *SERPINA1*^17^. The transmembrane serine protease 6 (encoded by *TMPRSS6*) is a negative regulator of hepcidin^65^, a key hormone regulator of iron homeostasis^13^. Given the role of the transmembrane serine protease 6 in iron regulation, both the serine protease inhibitors could potentially affect iron regulation via this gene.

Using DEPICT, we detected an enriched expression of genes in iron status loci in the liver. This is in line with the important role of the liver in iron metabolism and storage^66^, including hepcidin production. Consistent with previous studies, our analysis prioritized genes encoding known hepcidin regulators such as *HFE, TMPRSS6, TF, TFRC, TFR2, ERFE* and *IL6R*^67–70^. Mutations in several of these genes have been demonstrated to cause diseases of iron deficiency or overload^1,15,33,71^. The associations with other genes related to inflammation, both in the DEPICT and colocalization analyses, could possibly be related to the hepcidin response to inflammation. The genes and gene sets prioritized by DEPICT pointed to several different biological processes, which might reflect the numerous roles of iron in the body. A limitation with using a similarity-based method for gene set and gene prioritization for iron traits was that the software excluded the MHC region from the analysis, thereby also excluding one of the most central genes in iron homeostasis, the *HFE* gene.

Colocalization analysis further linked the GWAS loci both to the liver and to iron overload: iron status loci overlapped with cis-eQTLs for several of the hepcidin regulators, and genes involved in other liver functions such as lipid and fatty acid metabolism (*ORMDL1, FADS1*)^72,73^. The latter was also found in previous studies^35^ and is in line with the results from the PheWAS analysis. The colocalization of iron status loci and cis-eQTLs were however found in several tissues, and not primarily in liver. A limitation to the analysis was however that there were different sample sizes for each tissue, where liver had a relatively small sample size and subsequently lower power than other tissues.

Because iron plays an essential role in so many biological processes, several Mendelian randomization studies have explored the causal effect of iron status on a range of diseases^18–23^. Despite the known harmful effects of both iron deficiency and overload, no previous MR studies have investigated the shape of the exposure-outcome relationship. We therefore assessed the causal effect of iron status biomarkers on all-cause mortality and investigated the shape of these associations. We demonstrated that the GRSs based on the previous study were good instruments for iron, TSP and TIBC in the independent HUNT study (variance explained 6% (iron), 11% (TSP), 13% (TIBC), 1.6% (ferritin)). Using these, we found evidence of a harmful effect of increased serum iron and TSP (derived from serum iron and TIBC) in linear analyses that estimated population-averaged effects. The point estimates of TIBC and ferritin were also suggestive of a harmful effect of increased iron status, although the estimates were not statistically significant, and the ferritin estimate was very imprecise due to the small sample size. In non-linear models, we did not find strong statistical evidence supporting non-linear relationships over linear ones. However, there was weak evidence of a protective effect of increasing serum iron at the very low end of its distribution, at serum iron levels below the normal range of 10-34 µmol/L^51^. The results were supported by post-hoc sensitivity analyses using only genetic variants consistent with systemic iron status.

The study had several clear limitations. First, in our GWAS analyses we did not adjust for additional factors that could affect the biomarker concentrations, such as iron supplementation, inflammatory status, alcohol consumption and menopausal status (except for ferritin, where the full sample was pre-menopausal). These factors could therefore have influenced the effect estimates, particularly for rare variants. Second, we would need a larger sample size to confirm a non-linear shape of the exposure-outcome relationship at the extremes of the biomarker distributions. The analysis of ferritin was particularly limited by the small sample (N=2334) consisting of only relatively young, non-pregnant females, giving a low number of strata and few deaths. Third, the association of the GRSs with all-cause mortality could be attenuated because HUNT participants with suspected iron deficiency anemia or phenotypic hemochromatosis were later contacted by the primary health care services and offered treatment, and they could therefore have obtained a healthier iron status than they would otherwise have had, causing the analysis to be less precise. Finally, although the four biomarkers are commonly used to assess people’s iron status, neither of them is a very good individual predictor of iron stores, and the findings should therefore be interpreted with caution.

In summary, we have increased the number of iron status associated loci through a large GWAS meta-analysis and validated the latest genetic risk scores for four iron status biomarkers. We find evidence of a harmful population-averaged effect of genetically proxied serum iron and TSP, and weak evidence of a protective effect of increasing serum iron in individuals at the very low end of its distribution. Our findings contribute to our understanding of the genes affecting iron status and its consequences on human health.

## Methods

### Cohort descriptions

Distributions of the age and sex of the HUNT, MGI and SardiNIA participants included in the current study are reported in Supplemental Table 2.

#### HUNT

The HUNT Study is a longitudinal population-based health study conducted in the county of Trøndelag, Norway since 1984^25^. About 123 000 individuals (aged ≥20 years) have participated in at least one of four surveys, and more than 70 000 of these participants have been genotyped using one of three Illumina HumanCoreExome arrays: 12 v.1.0, 12 v.1.1 and 24 with custom content (UM HUNT Biobank v1.0). Sample and variant quality control (QC) was performed using standard practices and has been reported in detail elsewhere^74^. All variants were imputed from the TOPMed reference panel (freeze 5)^24^ using Minimac4 v1.0 (https://genome.sph.umich.edu/wiki/Minimac4). The reference panel is composed of 53 831 multi-ethnic samples and 410 323 831 SNP and indel variants at high depth (mean read depth 38.2X). Variants with a minor allele count (MAC) > 10 or imputation R^2^ ≥0.3 were included in analysis. A subset of individuals was genotyped with additional custom content variants.

### MGI

The Michigan Genomics Initiative (MGI) is a repository of genetic data and electronic medical records from Michigan Medicine. Approximately 80 000 participants (aged ≥18 years) have predominantly been enrolled prior to surgical procedures with over 59 000 individuals genotyped using Illumina Infinium CoreExome-24. Following genotyping, sample-level QC was performed to remove sex-mismatches, duplicates, samples with call rate < 99%, or with estimated contamination > 2.5%. Variants with GenTrain score < 0.15, Cluster Separation scores < 0.3, Hardy-Weinberg Equilibrium p-value among unrelated European individuals < 1×10^−4^, or with evidence of batch effects (p-value < 1×10^−3^, Fisher’s exact test) were excluded. Imputation was performed using the TOPMed Imputation Server (v1.2.7). Variants with MAF > 0.05% and imputation quality R^2^≥0.3 were included in analysis.

### SardiNIA

The SardiNIA study is a longitudinal population-based health study including 6 602 individuals from the Lanusei valley on Sardinia. The participants have been genotyped on four different Illumina Infinium arrays, OmniExpress, Cardio-Metabochip^75^, Immunochip^76^ and Exome Chip), and then imputed from a SardiNIA specific sequencing panel (∼4x coverage) of 3 839 individuals, using Minimac3^77^. Markers with imputation quality R^2^ > 0.3 (or > 0.6 in variants with MAF < 1%) were retained, resulting in a total of ∼19 million genetic variants. Samples, genotyping, sequencing and variant calling have previously been described elsewhere^78^.

### Iron Status Biomarkers

Distributions of the biomarker levels in the HUNT, MGI and SardiNIA participants included in the current study are reported in Supplemental Table 2.

#### HUNT

Non-fasting serum samples were drawn in 1995-1997 (HUNT2). Serum iron concentration (µmol/L) was determined using a FerroZine method using a Hitachi 911 Autoanalyzer (reagents from Boehringer, Germany). The serum transferrin concentration (µmol/L) was analyzed by an immunoturbidimetric method using the Hitachi 911 Autoanalyzer (reagents from DAKO A/S, Denmark), and calculated for a molecular weight of 79 570 Da. TIBC was calculated as 2 x the serum transferrin concentration. The TSP was calculated as 100 x [serum iron concentration/TIBC]. Serum ferritin was measured from serum samples using an Abbott AxSYM analyzer (reagents from Abbott Laboratories, USA). In total, 56 667 HUNT participants had measurements of serum iron and TIBC, 56 664 had measurements of TSP, while ferritin was only measured in 2 334 women (fertile, non-pregnant, aged 20-55 years).

#### SardiNIA

Serum iron (µmol/L) and serum transferrin concentrations (µmol/L) were measured in fasting blood-samples from individuals with genotype and imputation data from the SardiNIA cohort. TIBC was calculated as 2 x the serum transferrin concentration. In total, 5 930 and 5 926 genotyped SardiNIA participants had measurements on serum iron and TIBC respectively.

#### MGI

Serum iron concentration was measured using the Ferrozine Colorimetric assay, and serum ferritin was measured using a Chemiluminescent Immunoassay. Serum transferrin concentrations were measured using an Immunoturbidimetric assay, and TIBC was calculated as 2 x the serum transferrin concentration. The TSP was calculated as 100 x [serum iron concentration/TIBC]. For individuals with multiple measurements, the initial measurement was used in the analyses. In total, 10 403, 9 480, 10 399 and 10 381 participants from MGI had measurements of serum iron, serum ferritin, TIBC and TSP respectively.

### Association analyses

Association analyses of all iron traits (iron, ferritin, TIBC, TSP) in HUNT were performed using a linear mixed model regression under an additive genetic model for each variant as implemented in BOLT-LMM v2.3.4^79^, which also controls for relatedness between the samples. Association analyses of all iron traits in MGI were performed using a linear regression model in unrelated individuals using rvtests^80^. In both HUNT and MGI, we applied rank-based inverse normal transformation on the iron variables after adjusting for age and sex using linear regression, and included age, sex, genotyping batch and the first 10 principal components (PCs) of ancestry as covariates. Association analyses of serum iron and TIBC in SardiNIA was performed using age, age^2^ and sex-adjusted inverse-normalized residuals of TIBC or iron as input to the Efficient Mixed Model Association eXpedited (EMMAX)^81^ single variant test as implemented in EPACTS (https://github.com/statgen/EPACTS).

Additionally, we performed association analyses of serum iron, TIBC and TSP in HUNT with 361 965 additional custom content variants genotyped in 44 248 (serum iron, TIBC) or 44 246 individuals (TSP) using BOLT-LMM v2.3.4^79^, including the same covariates and rank-based normal transformation of the variables as was done in the main analyses.

### Meta-analyses

We performed fixed-effect inverse-variance weighted meta-analysis of summary statistics for iron (sample size N=236 612), ferritin (N=257 953), TIBC (N=208 422) and TSP (N=198 516) using METAL^82^. Serum iron and TIBC were meta-analyzed in all studies (HUNT, MGI, SardiNIA and summary statistics from deCODE and Interval). SardiNIA did not have data on serum ferritin and TSP and was therefore excluded from these meta-analyses, while the available summary statistics for ferritin also included the DBDS study. To harmonize genomic positions from each study, we used LiftOver from UCSC^83^ to map the data from SardiNIA from Human Genome Build GRCh37 to GRCh38. Because standard errors were not given in the available summary statistics from deCODE, Interval and DBDS, we calculated them as the absolute value of the (effect size/qnorm(p-value/2)), where qnorm represents the inverse standard normal distribution. In HUNT, MGI and SardiNIA we performed genomic control correction of any analyses with an inflation factor λ >1 prior to meta-analysis. We considered genetic loci reaching a p-value < 5×10^−8^ for follow-up analyses.

### Definition of independent loci and locus novelty

Genetic loci were defined around variants with a genome-wide significant association with a trait (p-value < 5×10^−8^). The locus borders were set 500 kb to each side of the highest and lowest genetic positions reaching genome-wide significance in each region. Overlapping genetic loci were merged if the index (lowest p-value) variants were in LD (R^2^>0.2 and/or D’≥0.8), or if one of the index variants was too rare to calculate LD with the other from our reference panel of 5000 unrelated individuals in HUNT. A locus was classified as novel for a given trait if it had not been reported previously for the trait. Previously published variants were identified through a literature search and a look-up in the GWAS catalog^84^.

### Annotation of genetic variants

We used plink v1.9^85^ with a reference panel of 5000 unrelated individuals in HUNT to identify genetic variants in strong LD (R^2^>0.8) with the index variants, and annotated the functional consequences and rsIDs of the index variants and LD proxies using ANNOVAR (v.2019Oct24)^86^ and the UCSC human genome browser^83^.

### Functional mapping of genetic variants

We used three different bioinformatic approaches to perform functional mapping and gene prioritization of the meta-analysis summary level data: Bayesian colocalization analysis^87,88^, DEPICT^47^ and Polygenic Priority Scores^89^.

To assess if any iron status loci were overlapping with significant cis-eQTL signals and consistent with shared causal variants for iron status markers and gene expression levels in specific tissue types, we used Bayesian colocalization analysis (‘coloc’) as implemented in the R package coloc. We used cis-eQTL data from 27 general tissue types (49 subtypes) in the individuals of European ancestry from the Genotype-Tissue Expression (GTEx) portal, data set v8 (https://www.gtexportal.org) and the GWAS meta-analysis results for each iron trait as input. For each tissue type, we analyzed all genes whose expression were associated (p-value < 5×10^−8^) with at least one iron status associated variant (p-value < 5×10^−8^), using effect sizes and standard errors for each variant-trait association as input. The coloc software estimated the variance in each trait (iron trait or gene expression level) from the sample sizes and minor allele frequencies. We set the prior probability of a genetic variant being associated with only iron traits, only gene expression or both traits to be 10^−4^, 10^−4^ and 10^−6^ respectively. We considered posterior probabilities (PP4) above 75% to give support for a common causal variant for the iron trait and expression of the gene in the given tissue.

We performed gene set enrichment, gene prioritization and tissue/cell type enrichment tests on the iron trait loci (p-value < 5×10^−8^) using Data-driven Expression Prioritized Integration for Complex Traits (DEPICT) (v1.1, release 194)^47^. Prior to the analysis we used LiftOver from the UCSC^83^ to convert the genomic positions of the genetic variants from GRCh38 to GRCh37. Enrichment results with an FDR < 5% were considered significant.

Finally, we prioritized genes by computing Polygenic Priority Scores^89^ from summary-level data from each iron status biomarker. The method uses Multi-marker Analysis of GenoMic Annotation (MAGMA)^90^ to compute gene-level associations and gene-gene correlations from the meta-analysis p-values and sample sizes and LD information from individuals of European ancestry from the 1000 Genomes reference panel^91^. MAGMA is applied a second time to perform enrichment analysis for genetic features. Genes are finally prioritized based on a combination of physical distance to associated genetic variants and functional similarity with other associated genes. We considered the 1% top ranked genes per biomarker to be prioritized genes for the respective traits.

### Heritability estimation

We estimated the narrow-sense additive SNP heritability of serum iron, TIBC and TSP in HUNT using GCTA^36^. Ferritin heritability was not estimated because of the low sample size in HUNT. We created genetic relationship matrices (GRMs) based on 358 956 genotyped autosomal variants in 56 667 individuals with serum iron and TIBC data, and 56 664 individuals with TSP data. We used the respective GRMs with GCTA-GREML (genomic-relatedness-based restricted maximum-likelihood) to estimate the variance in each variable that was explained by the genetic variants. We used age, sex, and genotyping batch as covariates in the analyses, and we transformed the iron trait variables to normality with rank based inverse normal transformation after regression on age and sex prior to the analyses.

### Genetic correlation between iron traits

We used the LDSC software^37^ with the iron trait meta-analysis summary statistics and precomputed LD Scores for Europeans from the 1000 Genomes reference panel^91^, and estimated the pair-wise genetic correlations of the four iron traits. Prior to the analysis, we changed all p-values < 1×10^−300^ to the exact value 1×10^−300^ to make sure the software was able to read the smallest values and did not discard these SNPs. To ensure that only well imputed SNPs were included in the analysis and thereby avoid bias due to variable imputation quality, we filtered the input files to the HapMap3 reference panel prior to the analysis, as recommended by the software developers (https://github.com/bulik/ldsc/).

### Phenome-wide association tests (PheWAS)

We constructed GRSs for the iron status biomarkers by summing the product of the effect size and the estimated allele count (dosage) for the index variants in genome-wide significant loci (p-value < ×10^−8^). We used TOPMed imputed estimated allele counts and effect sizes from the meta-analysis and calculated the GRS for participants of white British ancestry in the UK Biobank. We tested the association of the GRSs (GRS-PheWAS) and the individual index variants (single variant PheWAS) with 1 688 phecodes, continuous traits and blood biomarkers. We used a logistic or linear regression model respectively to assess the association of the single variant estimated allele counts (‘dosage’) or inverse normalized GRS and each phecode or continuous trait/biomarker. For the GRS-PheWAS we included as covariates sex and birth year for binary traits, and sex and age at measurement for continuous traits. For the single variant PheWAS we used GWAS summary statistics generated with SAIGE v.29.4.2^92^, with sex and the first four PCs as covariates in addition to age at initial assessment for quantitative traits and birth year for binary traits. To correct for multiple testing, we used a Bonferroni corrected p-value significance cut-off of 2.4×10^−7^, correcting for the number of tested variants, phecodes, biomarkers and continuous traits. In total, 29 variants were excluded from the single variant PheWAS and GRS-PheWAS because they were not imputed in UK Biobank (Supplemental Information, Section II)

### Validation of genetic risk scores in HUNT

To validate the previously published results from Iceland, Denmark and UK, we created weighted GRSs for each trait based on the published index variants and effect sizes^17^ using the same approach as described in the previous section. We tested the predictive ability of each GRS by regressing each trait on the respective GRS in the independent cohort, HUNT (N_iron_=N_TIBC_=56 667, N_TSP_=56 664, N_ferritin_=2334) and report the trait variance explained by the GRS. In total, ten variants were excluded from the GRSs because they were not imputed in HUNT (Supplemental Information, section III).

### Mendelian randomization of iron status on all-cause mortality

To assess the causal association of iron status on all-cause mortality, we performed linear MR analyses using the ratio of coefficients method^93^, using GRSs as genetic instruments for the four iron related traits. The GRSs were constructed as described above, using index variants and external effect sizes from the previous independent meta-analysis^17^. We used linear regression to estimate the associations between the iron related traits and the GRS, and a Cox proportional hazards regression to estimate the association of the GRS with mortality. The MR estimate was obtained as the ratio of the outcome-instrument and exposure-instrument association estimates. The standard error was estimated as the standard error of the GRS-mortality association divided by the GRS-biomarker association estimate.

To further assess the shape of the association, we performed a non-linear Mendelian randomization with the fractional polynomial method^94,95^ in HUNT, using the same GRS as genetic instrument for the iron traits. The method has been described in detail elsewhere^95–98^. In brief, each iron related trait was regressed on its respective GRS, and the population was divided into 100 (iron, TIBC, TSP) or 20 (ferritin) strata based on the residual traits. The number of strata was reduced for ferritin because of the lower sample size for this biomarker. In each stratum we used linear regression to estimate the association of the GRS with the iron trait, and Cox proportional hazards regression to estimate the association of the GRS with mortality. We calculated the localized average causal effect (LACE) of the respective trait on all-cause mortality in each stratum as the ratio of the GRS-outcome and GRS-exposure associations. We performed meta-regression of the LACE against the mean of the exposure in each stratum and tested whether the best-fitting fractional polynomial of degree 1 fitted the LACE estimates better than a linear model using the fractional polynomial method^94^.

To further validate the selection of SNPs representing each biomarker in the non-linear MR, we performed post-hoc sensitivity analyses rerunning the non-linear MR method with new instruments that had stricter criteria for SNP inclusion. Here, we restricted the GRSs to index variants from the previous study^17^ that were not only GWAS significant for at least one trait, but also nominally significant (p-value<0.05) for the remaining traits. Further, we excluded SNPs that had directions of effect that were not consistent with systemic iron status (increasing serum iron, ferritin and TSP, and decreasing TIBC)^99^. We used the remaining 14 SNPs (Supplemental Information, Section IV) to construct each of the four GRSs in the analysis as described for the main analysis.

### Ethics

All study participants have given informed consent. The analyses in HUNT has approval from the Norwegian Data Protection Authority and the Regional Ethics Committee for Medical and Health Research Ethics in Central Norway (REK Reference Number: 2014/144), the analyses in MGI are approved by the Institutional Review Board of the University of Michigan Medical School (IRB Reference Number: HUM00094409), and the analyses in UK Biobank are covered by the ethics approval for UK Biobank studies (application 24460) from the NHS National Research Ethics Service on 17th June 2011 (Ref 11/NW/0382) and extended on 10th May 2016 (Ref 16/NW/0274).

## Data Availability

Data generated or analyzed during this study are available from the corresponding authors upon reasonable request.

## Acknowledgements

The Trøndelag Health Study (The HUNT Study) is a collaboration between HUNT Research Center (Faculty of Medicine and Health Sciences, NTNU, Norwegian University of Science and Technology), Trøndelag County Council, Central Norway Regional Health Authority, and the Norwegian Institute of Public Health. The genotyping in HUNT was financed by the National Institutes of Health; University of Michigan; the Research Council of Norway; the Liaison Committee for Education, Research, and Innovation in Central Norway; and the Joint Research Committee between St Olav’s hospital and the Faculty of Medicine and Health Sciences, NTNU. The authors acknowledge the Michigan Genomics Initiative participants, Precision Health at the University of Michigan, the University of Michigan Medical School Central Biorepository, and the University of Michigan Advanced Genomics Core for providing data and specimen storage, management, processing, and distribution services, and the Center for Statistical Genetics in the Department of Biostatistics at the School of Public Health for genotype data curation, imputation, and management in support of the research reported in this publication.

## Author Contributions

M.R.M analyzed the data and wrote the first draft of the manuscript. A.F.H., S.E.G, S.A.G.T, K-H.W and W.Z. contributed to analyses. K.T. gave advice on phenotype definitions and hemochromatosis. S.B., A.M. and D.G. gave advice on non-linear Mendelian randomization. B.M.B, J.B.N, K.H. and C.J.W conceived and designed the study. All authors interpreted results and revised the paper.

