## Supplemental Information for "Genome-wide meta-analysis of iron status biomarkers and the effect of iron on all-cause mortality in HUNT"

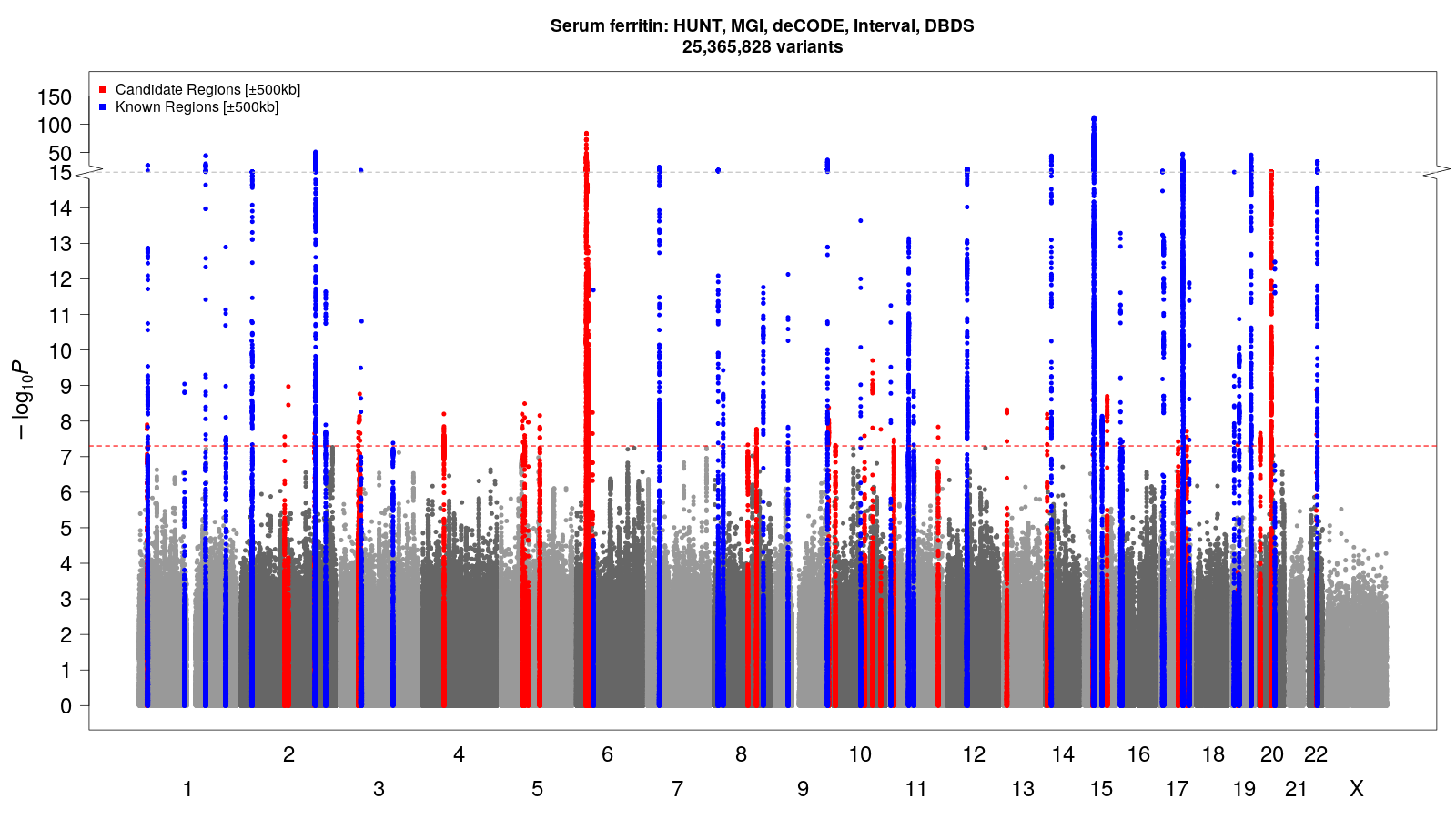

Supplemental Figure 1: Manhattan plot of the serum ferritin meta-analysis of the HUNT, MGI, deCODE, DBDS and Interval studies: The x-axis gives the chromosomes and chromosomal positions, and the y-axis gives -log10(p-value) for the genetic variants.

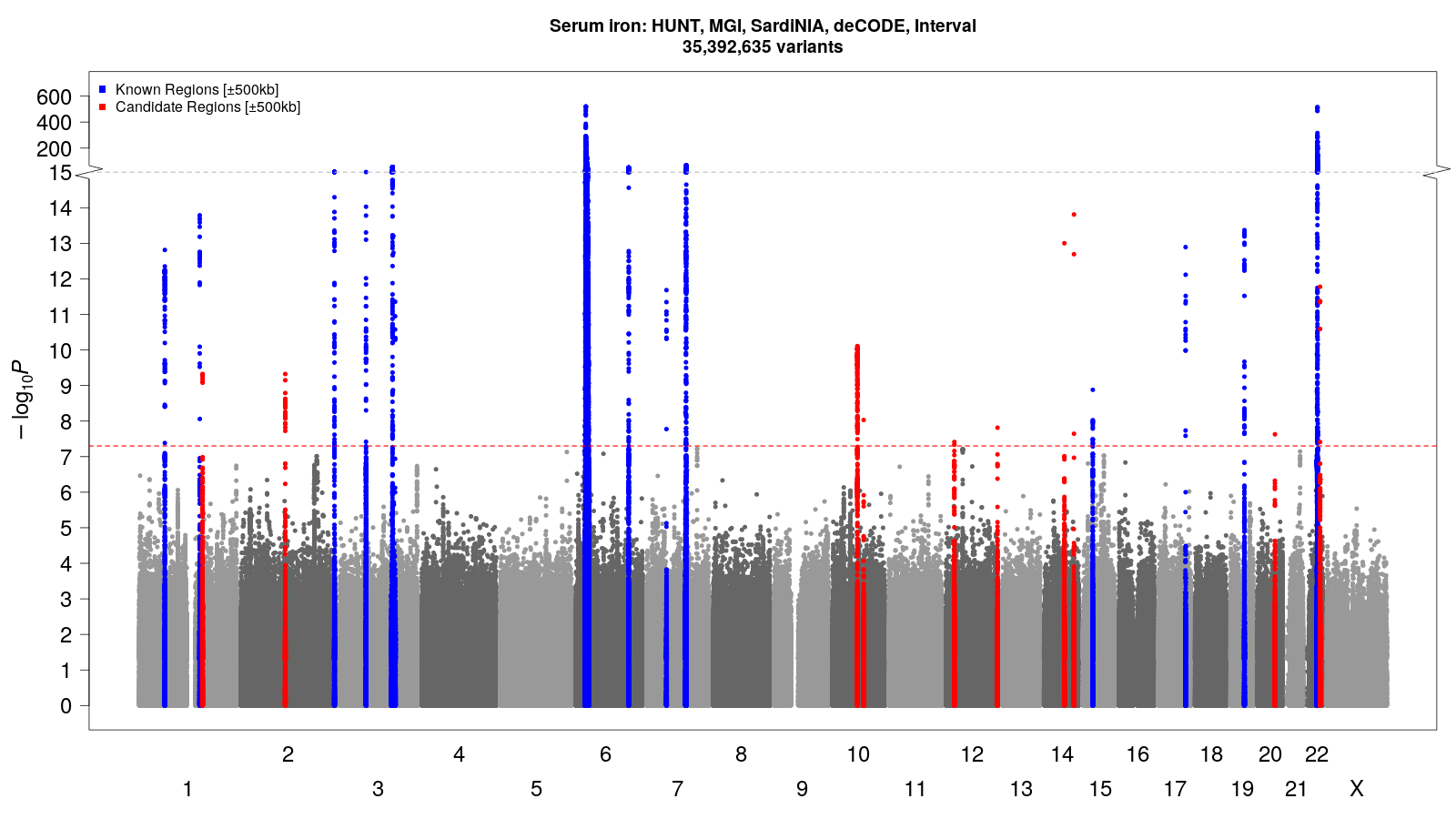

Supplemental Figure 2: Manhattan plot of the serum iron meta-analysis of the HUNT, MGI, SardiNIA, deCODE and Interval studies: The x-axis gives the chromosomes and chromosomal positions, and the y-axis gives -log10(p-value) for the genetic variants.

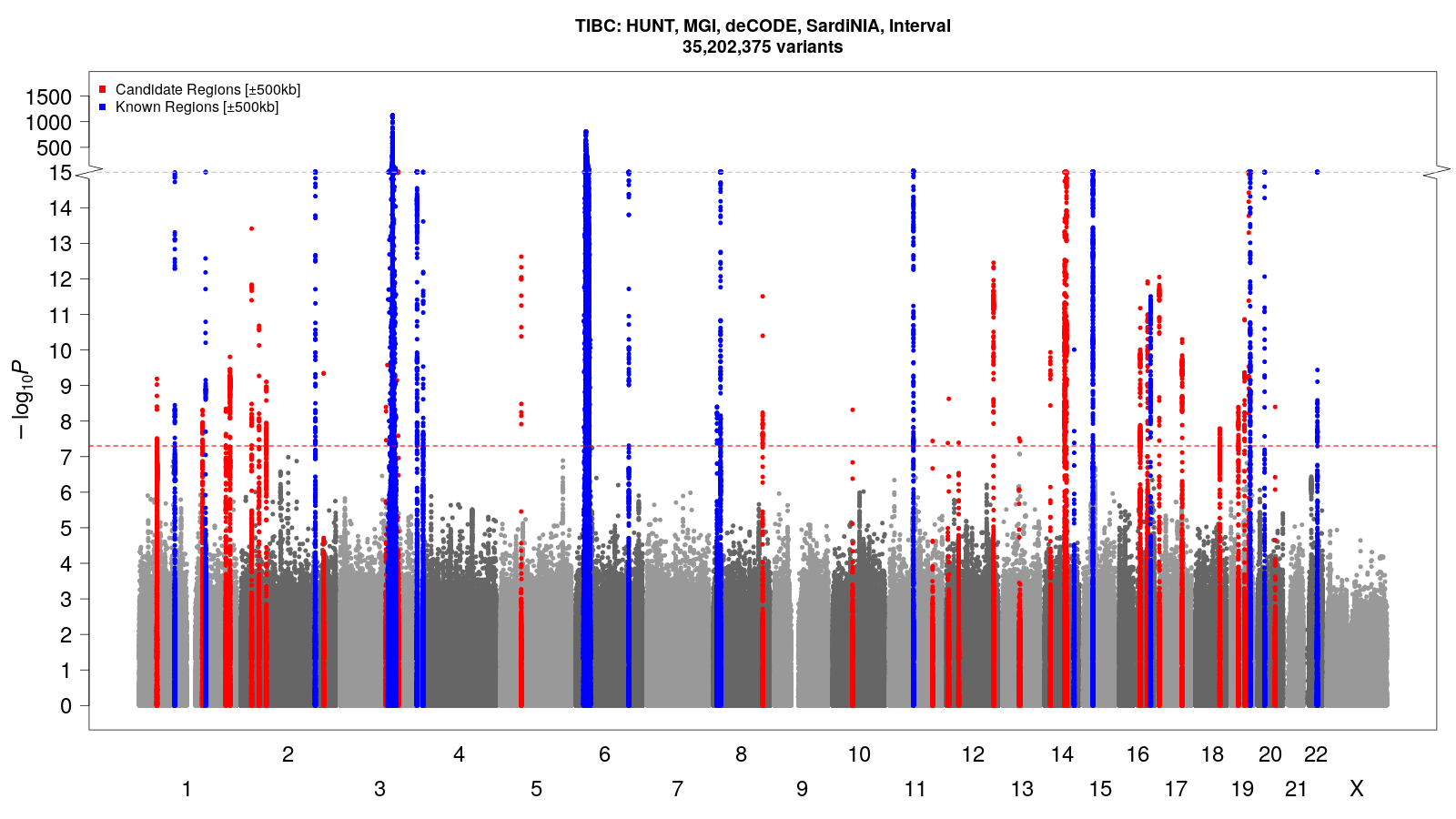

Supplemental Figure 3: Manhattan plot of the total iron binding capacity (TIBC) meta-analysis of the HUNT, MGI, SardiNIA, deCODE and Interval studies: The x-axis gives the chromosomes and chromosomal positions, and the y-axis gives -log10(p-value) for the genetic variants.

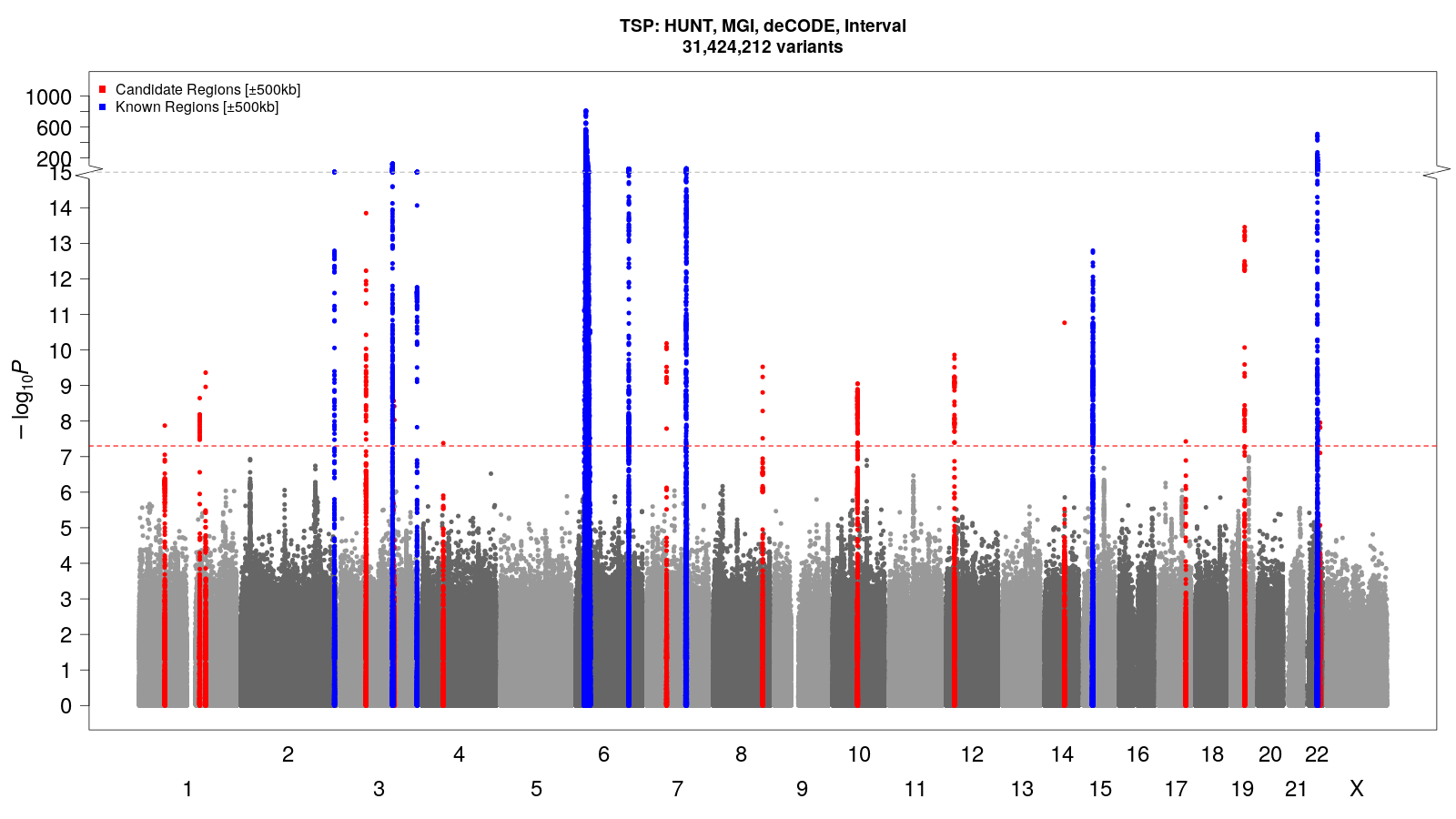

Supplemental Figure 4: Manhattan plot of the serum iron meta-analysis of the HUNT, MGI, deCODE and Interval studies: The x-axis gives the chromosomes and chromosomal positions, and the y-axis gives -log10(p-value) for the genetic variants.

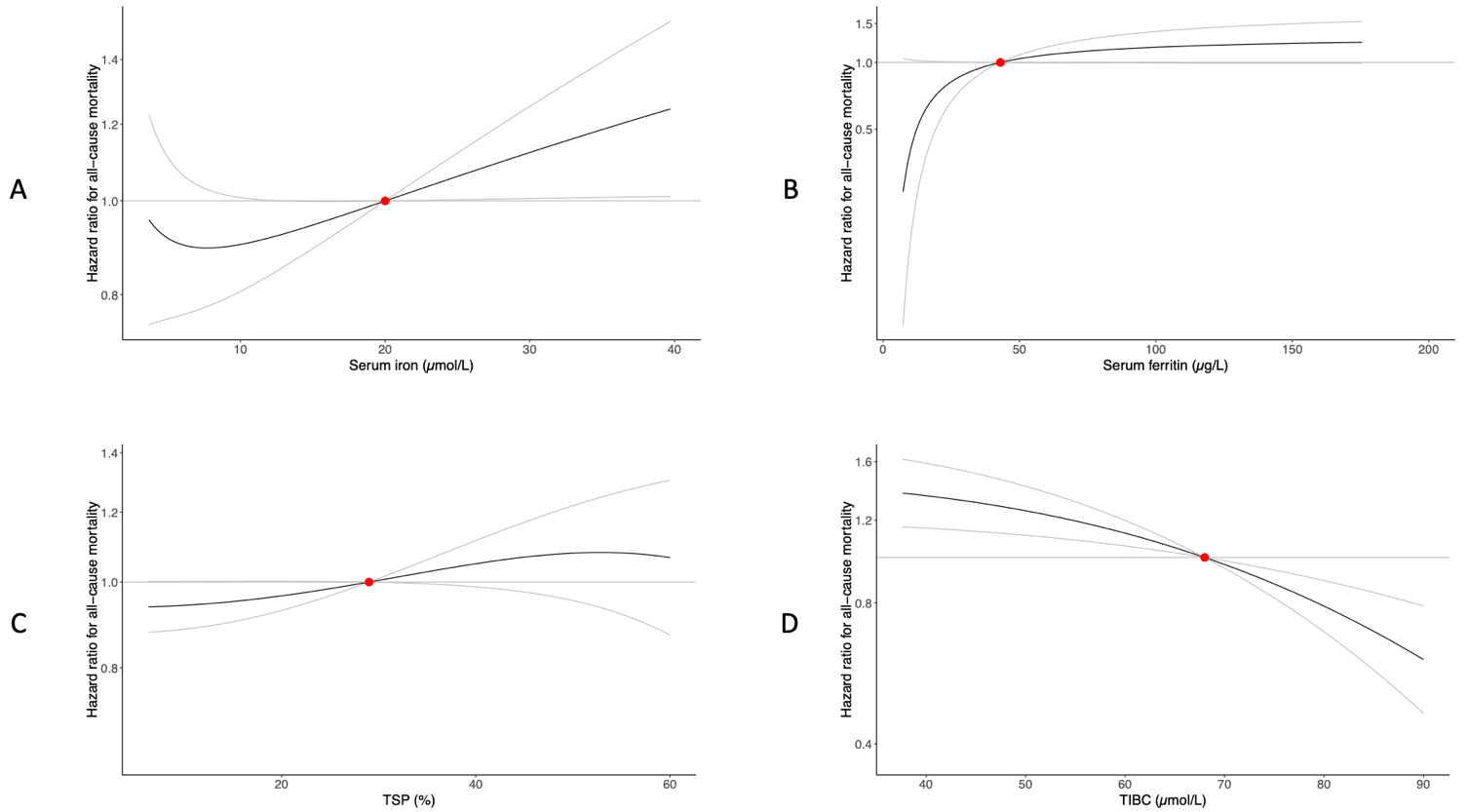

Supplemental Figure 5: Non-linear Mendelian randomization sensitivity analysis: Dose-response curves (black) between iron traits and all-cause mortality in HUNT (gray lines give 95% confidence interval) using a GRS consistent with systemic iron status. The x-axis gives A: serum iron levels (µmol/L), B: serum ferritin (µg/L), C: transferrin saturation (%) and D: total iron binding capacity (TIBC) (µmol/L). The y-axis gives the hazard ratios for all-cause mortality with respect to the reference values (red dot), which represent the established target values (iron, TIBC, TSP)^51^ or median value (ferritin) for the traits. The curve gradients represent the localized average causal effect at each point.

II) Genetic variants excluded from GRS-PheWAS because they were not imputed in the UK Biobank: rs61804206, rs374974760 (iron), rs10740134, rs35945185, rs2228145 (iron, TSP), rs7165401 (TSP), rs514595, rs469721, rs61830291, rs1435167, rs41274050 (TIBC), rs477190, rs79052526, rs10801913, rs551459670, rs142350264, rs536826368, rs7009973, rs189899297, rs681099, 9:133264504:G:GAAACTGCC, rs9423600, rs17476364, rs704017, rs556393026, rs10685744, rs141253118, rs192331981, rs608374 (ferritin).
III) Genetic variants excluded from the GRSs validation because they were not imputed in HUNT: rs35945185 (iron), rs748587164 (iron, TSP, TIBC), rs773570300 (iron, TSP), rs551459670 (ferritin), rs762752083 (ferritin), rs750717575 (ferritin), rs745795585 (ferritin), rs143041401 (ferritin) and two deletions on chromosomes 9 and 12 (ferritin).
IV) Genetic variants selected for GRS used in post-hoc sensitivity non-linear Mendelian randomization analyses: rs75965181, rs10801913, rs6025, rs13007705, rs1799945, rs1800562, rs9399136, rs4841429, rs13253974, rs2954029, rs57659670, rs34523089, rs2005682, rs855791. The directions of effect for these genetic variants were consistent with systemic iron status (increasing iron, TSP, ferritin, decreasing TIBC). Further, they were GWAS significant for at least one iron related biomarker and at least nominally significant for the other three biomarkers.
